## Supplementary material for "“The Running Injury Continuum: A qualitative examination of recreational runners’ description and management of injury”": Supplementary Material - The Running Injury Continuum.docx

**Supplementary material A: Focus group schedule, introduction & aims**

| **Domain** | **Sample dialogue** |
| --- | --- |
| Introduction & aims of study | Hi everyone. Thank you for coming and for being involved in this study. I am conducting some research on runners’ description and management of injury, and the aim of this study is to gather your thoughts on running-related injuries. Please go into as much detail as you can, ask each other questions, and agree or disagree on any points raised, but please respect everyone’s opinion. If you have any questions, please ask at any point. |
| Sample questions | How would you define injury? |
|  | How would you describe injury? |
|  | Based on your descriptions of injury and the terms you have used, could you elaborate on these on the whiteboard? |
|  | How would you manage injuries? |

**Supplementary material B: Standards for Reporting Qualitative Research (SRQR) Checklist (43)**

| **Topic** | **Page No(s).** |
| --- | --- |
| **Title and Abstract** | |
| **Title** - Concise description of the nature and topic of the study. Identifying the study as qualitative or indicating the approach (e.g., ethnography, grounded theory) or data collection methods (e.g., interview, focus group) is recommended | 2 |
| **Abstract** - Summary of key elements of the study using the abstract format of the intended publication; typically includes background, purpose, methods, results, and conclusions | 2 |
| **Introduction** | |
| **Problem formulation -** Description and significance of the problem/phenomenon studied; review of relevant theory and empirical work; problem statement | 3-5 |
| **Purpose or research question -** Purpose of the study and specific objectives or questions | 5 |
| **Methods** | |
| **Qualitative approach and research paradigm -** Qualitative approach (e.g., ethnography, grounded theory, case study, phenomenology, narrative research) and guiding theory if appropriate; identifying the research paradigm (e.g., postpositivist, constructivist/ interpretivist) is also recommended; rationale** | 6 |
| **Researcher characteristics and reflexivity -** Researchers’ characteristics that may influence the research, including personal attributes, qualifications/experience, relationship with participants, assumptions, and/or presuppositions; potential or actual interaction between researchers’ characteristics and the research questions, approach, methods, results, and/or transferability | - |
| **Context** - Setting/site and salient contextual factors; rationale** | 6 |
| **Sampling strategy –** How and why research participants, documents, or events were selected; criteria for deciding when no further sampling was necessary (e.g., sampling saturation); rationale** | 6-7 |
| **Ethical issues pertaining to human subjects -** Documentation of approval by an appropriate ethics review board and participant consent, or explanation for lack thereof; other confidentiality and data security issues | 6 |
| **Data collection methods -** Types of data collected; details of data collection procedures including (as appropriate) start and stop dates of data collection and analysis, iterative process, triangulation of sources/methods, and modification of procedures in response to evolving study findings; rationale** | 7-8 |
| **Data collection instruments and technologies -** Description of instruments (e.g., interview guides, questionnaires) and devices (e.g., audio recorders) used for data collection; if/how the instrument(s) changed over the course of the study | 7-8 |
| **Units of study -** Number and relevant characteristics of participants, documents, or events included in the study; level of participation (could be reported in results) | 11-12 |
| **Data processing -** Methods for processing data prior to and during analysis, including transcription, data entry, data management and security, verification of data integrity, data coding, and anonymization/de-identification of excerpts | 8-9 |
| **Data analysis -** Process by which inferences, themes, etc., were identified and developed, including the researchers involved in data analysis; usually references a specific paradigm or approach; rationale** | 8-9 |
| **Techniques to enhance trustworthiness -** Techniques to enhance  trustworthiness and credibility of data analysis (e.g., member checking, audit trail, triangulation); rationale** | 10-11 |
| **Results/Findings** | |
| **Synthesis and interpretation –** Main findings (e.g., interpretations, inferences, and themes); might include development of a theory or model, or integration with prior research or theory | 11-25 |
| **Links to empirical data –** Evidence (e.g., quotes, field notes, text excerpts, photographs) to substantiate analytic findings | 15-25 |
| **Discussion** | |
| **Integration with prior work, implications, transferability, and contribution(s) to the field -** Short summary of main findings; explanation of how findings and conclusions connect to, support, elaborate on, or challenge conclusions of earlier scholarship; discussion of scope of application/generalizability; identification of unique contribution(s) to scholarship in a discipline or field | 25-33 |
| **Limitations –** Trustworthiness and limitations of findings | 33-34 |
| **Other** | |
| **Conflicts of interest –** Potential sources of influence or perceived influence on study conduct and conclusions; how these were managed | - |
| **Funding –** Sources of funding and other support; role of funders in data collection, interpretation, and reporting | - |

**Supplementary material C: Order of themes:**

Table 1: Description and management of each level of the Running Injury Continuum

|  | **Core category** | **Theme** | **Sub-theme** | **Secondary sub-theme** | **Tertiary sub-theme** |
| --- | --- | --- | --- | --- | --- |
| **1** | **Running smooth** | | | | |
|  | Physical description | Sensation | Pain free |  |  |
|  | Management | Injury prevention |  |  |  |
|  |  | No management required |  |  |  |
|  | Psychological description | Happy place |  |  |  |
| **2** | **Discomfort** | | | | |
|  | Physical description | Sensation | Tightness |  |  |
|  |  |  | DOMS |  |  |
|  |  |  | Tiredness |  |  |
|  |  |  | Low pain |  |  |
|  |  |  | Stiffness |  |  |
|  |  |  | Uncomfortable |  |  |
|  |  | Frequency/Onset | Temporary |  |  |
|  |  |  | Infrequent |  |  |
|  |  | Precursor to injury |  |  |  |
|  | Outcome | Effect on performance | Full training | Stick to training plan |  |
|  |  | Effect on daily life |  |  |  |
|  | Psychological description | Mental fatigue |  |  |  |
|  | Management | Self-management | Additional stretching |  |  |
|  |  |  | Continue training |  |  |
|  |  |  | Therapies | Massage gun |  |
|  |  | External management | Friends |  |  |
|  |  |  | Internet sources | Google |  |
|  |  | No management |  |  |  |
| **3** | **Niggle** | | | | |
|  | Physical description | Sensation | Pain | Severity of pain | Low pain |
|  |  |  |  |  | Not painful |
|  |  |  |  |  | Dull pain |
|  |  |  |  | Type of pain | Aches |
|  |  |  | Awareness | Different from opposite side |  |
|  |  |  | Discomfort |  |  |
|  |  |  | Tightness |  |  |
|  |  |  | Irritation |  |  |
|  |  |  | Tiredness |  |  |
|  |  | Frequency/Onset | Repeated |  |  |
|  |  |  | Temporary |  |  |
|  |  |  | Constant |  |  |
|  |  |  | Post-session complaint |  |  |
|  |  |  | Felt while running |  |  |
|  |  | Precursor to injury | From an unknown cause |  |  |
|  | Outcome | Effect on performance | Full training | Run through it |  |
|  |  |  |  | Pressure to continue | Desire to continue |
|  |  |  |  |  | Stick to training plan |
|  |  |  |  | Doesn’t affect training |  |
|  |  |  |  | Can ignore |  |
|  |  |  | Altered training | Altered warm up |  |
|  |  |  |  | Forced change to training | Reduce load |
|  |  |  |  | Altered technique |  |
|  |  | Effect on daily life | No effect |  |  |
|  | Psychological description | Caution |  |  |  |
|  |  | Affects motivation to train |  |  |  |
|  |  | Annoying |  |  |  |
|  | Management | Self-management | Altered training | Additional stretching |  |
|  |  |  |  | Rest | Additional rest day |
|  |  |  |  | Reduce load |  |
|  |  |  |  | Session preparation |  |
|  |  |  |  | Change technique |  |
|  |  |  | Therapies | Foam rolling |  |
|  |  |  | Strength & conditioning |  |  |
|  |  |  | Accessory supports | Footwear |  |
|  |  | External management | Friends |  |  |
|  |  |  | Internet sources | YouTube exercises |  |
|  |  |  | Massage |  |  |
|  |  | No management | Resolves on its own |  |  |
|  |  |  | Ignore niggle |  |  |
|  |  |  | Ignore advice |  |  |
| **4** | **Twinge** | | | | |
|  | Physical description | Sensation | Pain | Type of pain | Darting pain |
|  |  | Frequency/Onset | Temporary (short-lived) |  |  |
|  | Outcome | Effect on performance | Altered training | Stop mid-session |  |
|  |  |  |  | Reduced load |  |
|  |  |  | Full training | Run through it |  |
|  | Psychological description | Caution |  |  |  |
|  |  | Annoying |  |  |  |
|  | Management | Self-management | Altered training | Reduce load |  |
|  |  |  |  | Rest | Stop mid-session |
|  |  |  |  | Additional stretching |  |
|  |  | External management | Friends |  |  |
|  |  |  | Google |  |  |
| **5** | **Persisting Niggle** | | | | |
|  | Physical description | Frequency/Onset | Persistent |  |  |
|  |  | Sensation | Pain | Severity of pain |  |
|  | Outcome | Effect on performance | Altered training | Reduce load |  |
|  |  |  |  | Rest days |  |
|  | Psychological description | Anxiety |  |  |  |
|  |  | Annoyed |  |  |  |
|  | Management | Self-management | Altered training | Reduce load |  |
|  |  |  |  | Stop running | Short-term rest |
|  |  |  | Therapies |  |  |
|  |  | External management | Friends |  |  |
|  |  |  | AT/Physio |  |  |
|  |  |  | Internet sources | Google |  |
| **6** | **Non-responsive niggle** | | | | |
|  | Physical description | Sensation | Pain | Pain stops running |  |
|  |  | Frequency/Onset | Constant |  |  |
|  |  |  | Pain while running |  |  |
|  | Outcome | Effect on performance | Altered training | Reduce load |  |
|  |  |  |  | Altered technique |  |
|  |  |  | Continue to train |  |  |
|  |  |  | Stop running | Short-term rest |  |
|  |  | Effect of daily life | Effects daily life | Pain during day |  |
|  |  |  |  | Conscious off-loading |  |
|  | Psychological description | Anxiety |  |  |  |
|  | Management | Self-management | Altered training | Stop running | Rest |
|  |  |  |  | Additional stretching |  |
|  |  |  |  | Reduce load |  |
|  |  |  | Medication |  |  |
|  |  | External management | AT/Physio |  |  |
|  |  |  | Friends |  |  |
|  |  |  | Failed self-management |  |  |
| **7** | **Injury: short term effect** | | | | |
|  | Physical description | Frequency/Onset | Progressively worsening |  |  |
|  |  |  | Acute onset | ‘Snap’ |  |
|  |  |  |  | ‘Pulled muscle’ |  |
|  |  | Sensation | Pain | Type of pain | Severe discomfort |
|  | Outcome | Effect on performance | Stop running | Unable to run - short term (weeks) |  |
|  |  |  | Continue to run (with pain) |  |  |
|  |  | Effect on daily life | Effects daily life | Pain during day |  |
|  |  |  |  | Conscious off-loading |  |
|  | Management | External management | AT/Physio | Stop running |  |
|  |  | Self-management | Rest |  |  |
|  | Psychological description | Anxiety |  |  |  |
| **8** | **Injury: long-term effect** | | | | |
|  | Physical description | Sensation | Pain | Extreme discomfort/Severe pain |  |
|  | Outcome | Effect on performance | Stop running | Unable to run - long term (months) |  |
|  |  | Effect on daily life | Effects daily life | Pain during day |  |
|  |  |  |  | Conscious off-loading |  |
|  | Management | External management | Medical specialist |  |  |
|  |  |  | AT/Physio |  |  |
|  |  | Self-management | Stop running |  |  |
|  | Psychological description | Anxiety |  |  |  |
|  |  | Frustration |  |  |  |
| **9** | **Career-ending injury** | | | | |
|  | Physical description | Frequency/Onset | Constant |  |  |
|  |  |  | Pan outside running |  |  |
|  |  | Sensation | Pain | Severity of pain | Very high |
|  | Outcome | Effect on performance | Unable to run - permanently |  |  |
|  |  |  | Career-ending injury |  |  |
|  |  | Effect on daily life | Effects daily life | Pain during day |  |
|  |  |  |  | Conscious off-loading |  |
|  | Management | External management | Requires surgery |  |  |
|  | Psychological description | Frustration |  |  |  |
|  |  | Depression |  |  |  |

parti: participants, FGs: focus groups, * indicates out of 31 participants, # indicates out of 7 participants

Table 2: Factors that influence runners’ description & management of the development of injury process

| **Core categories** | **Themes** | **Sub-themes** |
| --- | --- | --- |
| Running habits & history | Running experience |  |
|  | Motivations | Competitiveness |
|  |  | Chasing high |
|  |  | Goals |
|  |  | Stick to a plan |
|  | Other sport participation |  |
|  | Run setting | Group setting |
|  |  | Race |
|  |  | Individual |
|  |  | Training session |
|  | Event coming up |  |
|  | Knowledge |  |
| Individual | Individual perception |  |
|  | Daily life | Children |
|  |  | Mood |
|  |  | Menstrual cycle |
|  |  | Fatigue |
|  | Age |  |
|  | Sex |  |
| Injury | Previous injury |  |
|  | Type of injury |  |
